# Passive surveillance assesses compliance with COVID-19 behavioral restrictions in a rural US county

**DOI:** 10.1101/2020.09.09.20190389

**Authors:** Christina L Faust, Brian Lambert, Cale Kochenour, Anthony C. Robinson, Nita Bharti

**Affiliations:** Center for Infectious Disease Dynamics, Department of Biology, Eberly College of Science, Pennsylvania State University, University Park, PA, USA; GeoVISTA Center, Department of Geography, College of Earth and Mineral Sciences, Pennsylvania State University, University Park, PA, USA

**Keywords:** SARS-CoV-2, rural health, contacts, movement, behavioral restrictions

## Abstract

Following the emergence of SARS-CoV-2, early outbreak response relied on behavioral interventions. In the United States, local governments implemented restrictions aimed at reducing movements and contacts to limit viral transmission. In Pennsylvania, restrictions closed schools and businesses in the spring of 2020 and interventions eased later through the summer. In a rural Pennsylvania county, we use passive monitoring of vehicular traffic volume and mobile device derived visits to points of interest as proxies for movements and contacts. Rural areas have limited health care resources, which magnifies the importance of disease prevention. These data show the lowest levels of movement occurred during the strictest phase of restrictions, indicating high levels of compliance with behavioral intervention. We find that increases in movement correlated with increases in SARS-CoV-2 cases 9-18 days later. The methodology used in this study can be adapted to inform outbreak management strategies for other locations and future outbreaks that use behavioral interventions to reduce pathogen transmission.

## Introduction

Sudden Acute Respiratory Syndrome Coronavirus 2 (SARS-CoV-2), the virus that causes Coronavirus Disease 2019 (COVID-19) was first detected in Wuhan, China, in December 2019 [1]. The WHO declared COVID-19 was a pandemic in March 2020 (Figure 1) [2]. COVID-19 is a respiratory disease in humans and clinical presentation can include a variety of secondary symptoms, including gastrointestinal and neurological, that range from mild to severe or fatal [3,4]. The virus is spread by respiratory droplets, which can occur through close contacts [5]. Infectious individuals can be asymptomatic or mildly symptomatic [3]. Similar to early stages of other novel emerging pathogens, targeted pharmaceutical interventions for SARS-CoV-2 were limited. Large scale outbreak management efforts focused on behavioral interventions to reduce transmission [6–8]. Assessing changes in movement levels, contacts, and potential transmission events can help establish early warnings, implement adaptive control strategies, and disseminate preventative public health messaging to slow transmission.

**Figure 1.**
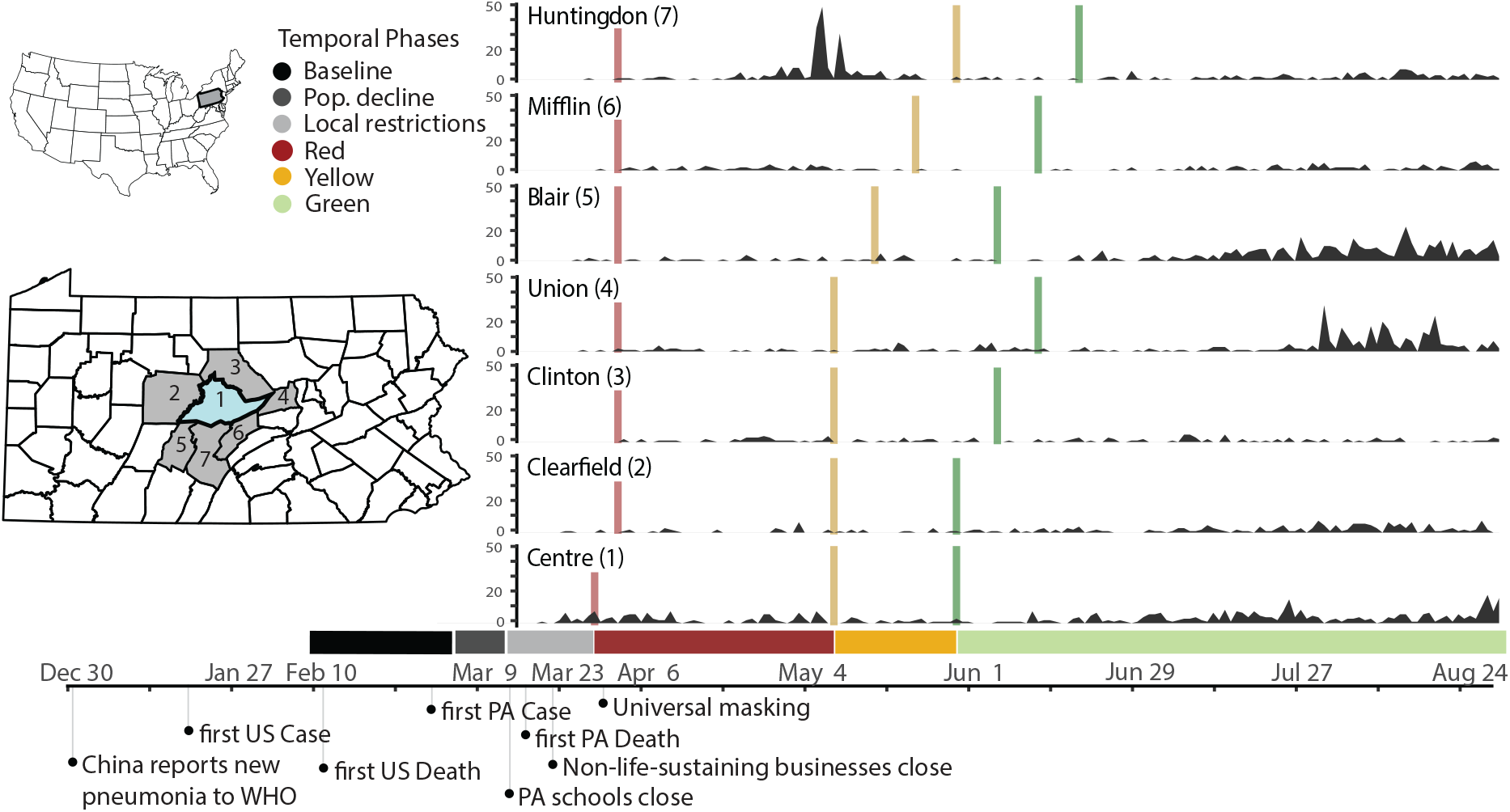
Timeline of policy interventions and cases in Centre County and adjacent counties. Left inset maps: location of Pennsylvania in the United States with locations of Centre County (blue) and surrounding counties (grey). Right: Daily case reports by county, color bars indicate onset of restriction phases. Below: Colors for all temporal phases for Centre County. Bottom: Timeline of relevant events.

There are many ways to measure human populations and movement to inform disease transmission. Data resolution varies across spatiotemporal scales, from targeted individual surveys and censuses [9] to large-scale passive surveillance data generated by satellites [10] and mobile devices [11]. Estimates of human movement and contacts for disease management have included tracking currency [12], commercial air traffic to model long distance flows [13], anthropogenic illumination to quantify seasonal or long-term population changes [10,14,15], and mobile devices for mobility traces [7,16,17]. Privately owned mobile device data are highly confidential and can’t be shared with policy makers. Researchers can obtain deidentified device data from third parties, which may be expensive or rely on opaque, proprietary algorithms. To overcome limitations on data sharing and increase replicability, we pair mobile device-derived data [18] with publicly available traffic camera images to measure human movement.

Early in the pandemic in the United States, county-level reporting showed that large cities and well-connected metro areas were hit the hardest. As the outbreak progressed, it characteristically moved into smaller towns and rural areas across the country [19]. Connectivity *between* locations loosely determines how early an infectious agent will arrive in a place and movement *within* a location determines how rapidly it can spread locally [20]. To reduce transmission, many states within the US introduced large scale quarantines targeting connectivity and movement. Understanding movement and pathogen transmission in rural areas is important because rural movement patterns differ from those found in urban areas [21,22] and residents in rural areas experience barriers to health care access that are different from barriers in urban areas [23]. Rural residents in the US have lower median household incomes and often rely on under-resourced health care services [24].

Centre County is located in rural valley in central Pennsylvania. It is home to The Pennsylvania State University’s (PSU) University Park (UP) campus, the largest campus of the state’s largest public institute of higher education. PSU is one of 112 land grant universities in the country. These institutions were established with a focus on agricultural education and improvement and, like PSU, many are still surrounded by suburban and rural areas. In 2019, the median US household income was $68,703 while the median Centre County, PA, household income was about 15% lower - $58,055 [25].

Prior to COVID-19, Centre County had 12 intensive care unit (ICU) beds, which increased to 24 during the pandemic [26]. Testing capacity remained severely limited throughout the US in 2020, including in Centre County. Test results were returned with delays of days or weeks, which hindered contract tracing efforts [27] and broad scale restrictions remained necessary.

Centre County, Pennsylvania, first implemented policies to minimize virus spread and transmission in March, in the form of travel restrictions and stay at home orders (Figure 1; Table 1). We monitored indicators of human moment in response to COVID-19 restrictions and the resulting outbreak trajectories from March through mid-August 2020, focusing on times when the university was not holding in-person classes and most students were not residing in the county. Our analysis begins six weeks prior to the strictest restrictions and ends during 2020’s most lenient county-level restrictions.

**Table 1.** Details of Pennsylvania COVID-19 Restriction Phases (dates for Centre County)

| **Restriction Phase** | **Length of phase** | **Work & congregate settings** | **Social settings** |
| --- | --- | --- | --- |
| Red | 40 days<br>(28 March– 7 May) | • Life-sustaining businesses only<br>• Schools closed for in-person teaching<br>• Most childcare closed | • Stay at home order<br>• Large gatherings prohibited<br>• Travel limited to life-sustaining purposes<br>• Restaurants/ bars limited to carry-out and delivery |
| Yellow | 20 days<br>(8 May – 28 May) | • Telework where feasible<br>• Businesses with in-person operations must follow safety guidelines<br>• Schools closed for in-person teaching<br>• Some childcare open with safety orders | • Full stay at home restrictions lifted<br>• In person retail allowed, but curbside/delivery preferred |
| Green | 78 days<br>(29 May– 14 August for this study; though restrictions | • Businesses reopen with CDC and PA Department of Health (DOH) guidelines (reduced | • Gyms, spas, entertainment reopen pending guidelines<br>• Aggressive mitigation orders lifted (individuals |
|  | were in place until<br>12 December) | capacity, distancing, mask<br>wearing) | must follow CDC and PA<br>DOH guidelines) |

We measured two proxies for movement within Centre County: vehicle volumes during the pandemic from publicly accessible traffic cameras and two years of mobile device derived visits to points of interest (POI) from SafeGraph [28]. Traffic volumes provide a good indication of movement in rural areas, which lack public transit and rely on private vehicles, but likely underrepresent movement in denser, pedestrian-heavy areas. Mobile device data likely under sample the elderly and residents in areas with poor network coverage or speed but are likely to adequately represent movement in the pedestrian-heavy areas on and surrounding university campuses. An advantage of SafeGraph’s ongoing passive surveillance is a long period of observation that precedes the perturbations caused by the pandemic.

Using these two data sources and official case reports, we compared movement across restriction phases to measure compliance and detect changes in confirmed SARS-CoV-2 cases. Overall, we found that movement measured by traffic volumes and mobile device derived visits to POI increased as restrictions eased, indicating compliance. We also found that cases increased as movements increased with a 9-to 18-day lag, suggesting that compliance with behavioral interventions was effective in reducing SARS-CoV-2 transmission.

The ongoing emergence of increasingly transmissible or virulent variants emphasizes the importance of accurately measuring the uptake of behavioral interventions and estimating the resulting impact on transmission dynamics. Our methods are broadly applicable across rural areas that are managing transmissible pathogens with behavioral interventions. The approaches presented here can inform outbreak management strategies on the effectiveness of strategies surrounding around behavioral interventions going forward.

## Methods

### Study area

Centre County has an estimated population size of 162,000 residents [25] and approximately 38,000 of these residents are undergraduate students enrolled at PSU’s UP campus [29]. Most undergraduate students do not reside in the county year-round, leading to declines in student population during the summer and winter breaks between semesters.

### Study period

We used traffic cameras and mobile device-derived visits to POI to detect changes in movement, with a focus on responses to restriction policies from 14 February 2020 to 14 August 2020 (Figure 1). We defined six temporal phases that align with events and policies that impacted movement and behavior at PSU’s UP campus and across Centre County in 2020 (Table 1, Figure 1):

1. **Baseline (14 February – 6 March)**: before restrictions were in effect and while undergraduate students were on campus
2. **Population Decline (7 March – 18 March)**: before local restrictions were in effect, after students left campus for spring break, and encompassing the transition to online instruction on March 16^th^
3. **Local Restrictions (19 March – 27 March)**: Mandated closure of all non-essential businesses (no county-wide restrictions)
4. **Red (28 March– 7 May)**: County-wide Red restriction phase
5. **Yellow (8 May 8 – 28 May)**: County-wide Yellow restriction phase
6. **Green (29 May 29 – 14 August)**: County-wide Green restriction phase

The calendar dates defining the Baseline (1), Local Restrictions (3), and Red (4) phases in 2020 occurred during the spring semester, which correspond to dates in 2019 when undergraduate students were in residence in Centre County. The Population Decline (2), Yellow (5), and Green (6) phases of 2020 coincided with times when semester classes were not in session in 2019 (spring break or summer session) and the student population in Centre County was drastically smaller. In 2020, students left campus during the Population Decline phase for spring break and were instructed not to return because the university implemented COVID-19 prevention policy to transition to online instruction from 16 March 2020.

To measure changes in movement through the restriction phases of 2020, we collected traffic data from 26 April 2020 to 14 August 2020. To measure changes in movement between years, we used SafeGraph mobile device derived counts of visits to POI from 14 February 2020 to 14 August 2020 and the corresponding periods in 2019 [28]. Pennsylvania began reporting confirmed SARS-CoV-2 cases on 1 March 2020 [30].

### Traffic Cameras

We collected images from 19 traffic cameras across Centre County to quantify numbers of vehicles on roads beginning on 26 April 2020 (Figure 2A). Twelve of these cameras surveil interstates, state highways, or other roads that link towns in Centre County, which we refer to as ‘connector’ roads (Table S1). The remaining seven cameras capture ‘internal’ roads for travel within towns. Cameras produce 24-hour live streams that are publicly accessible online but are not archived. We captured and stored images from these live streams approximately every 20 seconds. Using the Python package *cvlib* as a high-level interface to OpenCV [31,32] and Google’s open source TensorFlow software stack [33], we identified and counted all vehicles in each image. Parked vehicles were identified and excluded. The hourly number of vehicles captured by each camera was standardized by the number of images captured within that hour (see SM for details).

**Figure 2.**
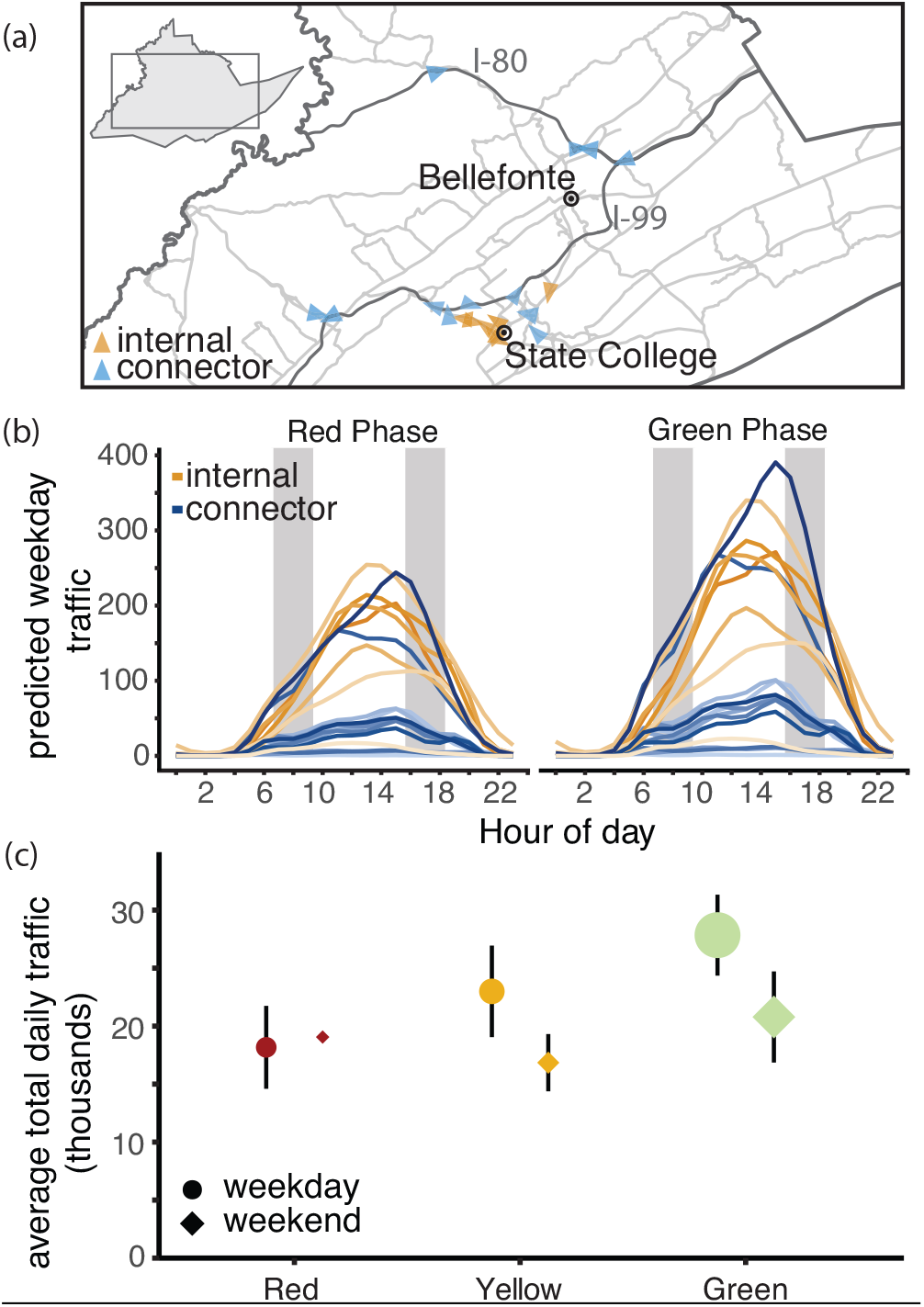
Traffic camera locations and volume throughout containment phases. (A) Locations of traffic cameras. Internal road traffic cameras (yellow) were concentrated around the PSU campus and surrounding boroughs, the connector road cameras (blue) covered a larger spatial extent. The direction of each arrow points to the direction the image was captured. (B) Centre County hourly vehicle volumes during Red and Green phases. Traffic during all phases peaked between 12:00 and 18:00 h EST. Vehicle volume was significantly greater in the green phase with the greatest increases on connector roads. Typical rush hours (07:00-09:00 and 16:00-18:00) are shaded in grey. (C) Traffic increased through each phase of easing of restrictions. The size of each point indicates the number of days of data collection. The smallest sample size collected was during weekends in the Red phase (2 days) and the largest number of observations was during weekdays in the Green phase (55 days).

We fit a series of generalized additive models (GAMs) to the standardized hourly vehicle counts. The effect of hour of the day in local time was modelled as a cyclic cubic regression spline to account for the continuity of the variable, meaning the same smoother operated at the 12:00 am ‘start’ of a day and the 11:00 pm ‘end’ of a day (Figure S1). We included the following predictor variables, with interactions between variables: day of week, weekday/weekend (binary variable), restriction phase (red/yellow/green), camera identity, number of lanes visible in camera image, and road type (internal or connector road). We fit multivariate GAMs using the package *mgcv* in R version 3.6.2 [34,35]. Most of the traffic cameras experienced at least one short gap in image acquisition (Figure S2). Missing hourly data were predicted using the best fit GAM. The combined data (both observed and predicted) were used to estimate vehicle traffic volumes and assess changes between restriction phases.

### Mobile device derived data

<u>SafeGraph</u>, a data company, provides aggregated anonymized location data from numerous applications in order to provide insights about physical places, here referred to as points of interests (POIs), via the <u>Placekey</u> Community. These data include mobile device derived counts of daily visits to POI (businesses, offices, university buildings, etc.). They sample visits from 45 million mobile devices across the United States to 3.6 million points of interest and their sampling correlates with census-derived population sizes [28]. In 2020, SafeGraph collected data in Centre County on 3,248,064 visit counts to 2,188 POI from a weekly mean of 5,165 mobile devices (range 2,993-7,831).

We used data collected in Centre County from 2019 and 2020. We used daily visit counts from early in 2020 before any COVID-19 restrictions were imposed, to establish expected visit counts during times when students were present and absent, in the Baseline and Population Decline phases, respectively. We used the differences to establish expected visits for comparison to observed visit counts while accounting for annual growth in SafeGraph data collection. These expectations help us quantify the changes in movements that were caused by the pandemic. To quantify the changes in movement through the restriction phases, we subtracted 2020 visit counts from 2019 visit counts. We aligned dates from 2019 to the corresponding academic periods in 2020 to account for differences in calendar dates (Table S1; [36]). We compared the differences in visit counts between years to the expected visit counts from early 2020 to quantify the observed different in visit counts due to behavioral interventions.

### COVID-19 diagnostic testing results

We acquired total daily confirmed cases of COVID-19 in Centre County and the surrounding counties from the Pennsylvania Department of Health’s version of the National Electronic Disease Surveillance System [30]. Cases are confirmed using CDC-approved diagnostic reverse transcriptase polymerase chain reaction (RT-PCR) tests, which detect viral RNA in active infections. The case data included here extend to 27 August 2020, two weeks past the last date of inclusion for movement data and transmission events in this study. We included this two week lag to account for a 5 to 8 day incubation period [37] and 5.5 to 10.93 day delay from the onset of symptoms to case confirmation [3]. We used cross correlation analyses to identify significant time-lagged correlations between movement measurements and the 7-day moving average of reported cases. We used the 7-day moving average of incidence because large numbers of positive test results reported on a single day may be due to tests that were processed in batches, which do not necessarily reflect a single-day increase in cases.

## Results

### Changes in traffic volume through restriction phases

Throughout restriction phases, from 26 April 2020 forward, daily traffic volume followed a bell-shaped pattern, with minimal traffic at 02:00 h and peaks between 12:00 and 18:00 h (Figure 2B, Figure S1). The best-fit Poisson GAM included splines fit to hourly average vehicle counts per camera, a binary weekend predictor, intervention phase, and an interaction between phase and road type (connector or local). This model explained 87.3% of deviance observed in vehicle traffic.

From April to August, Pennsylvania’s county-level behavioral restriction phases eased twice and traffic volume increased significantly. We observed greater increases in numbers of vehicles during weekdays and on connector roads than during weekends or on local roads (Figure 2B,C). During the Red phase, we calculated a daily weekday mean of 10,772 vehicles on internal roads and 7,397 vehicles on connector roads (Table S3). After Centre County eased to the Yellow phase of restrictions, mean daily weekday traffic totals increased by 23.2% on internal roads (an increase of 2,495 vehicles on weekdays) and 31.5% on connector roads (an increase of 2,331 vehicles on weekdays). When the county transitioned from the Yellow to the Green phase of restrictions, we saw an additional 15.4% increase in daily weekday traffic on internal roads (an increase of 2,042 vehicles on weekdays), and a 27.8% increase in vehicles on connector roads (an additional 2,697 vehicles on weekdays).

### Changes in mobile device derived visit counts between years and through 2020 restriction phases

Visit counts in 2020 were highest during the Baseline phase, when students were present (median daily visit count = 18,220) (Figure S3). When compared to the same period in 2019, there were more visits in 2020 (median difference = +4,613 in 2020)(Figure 3). Visit counts in 2020 began to decrease after the Baseline phase, during the Population Decline phase in March. This decline occurred when students departed for spring break and was matched by declines in 2019 (daily median differential of +464 visits in 2020). In 2019, visit counts increased immediately following spring break due to the return of students, while in 2020 the visit counts continued to decline due to the university’s pandemic protocol.

**Figure 3.**
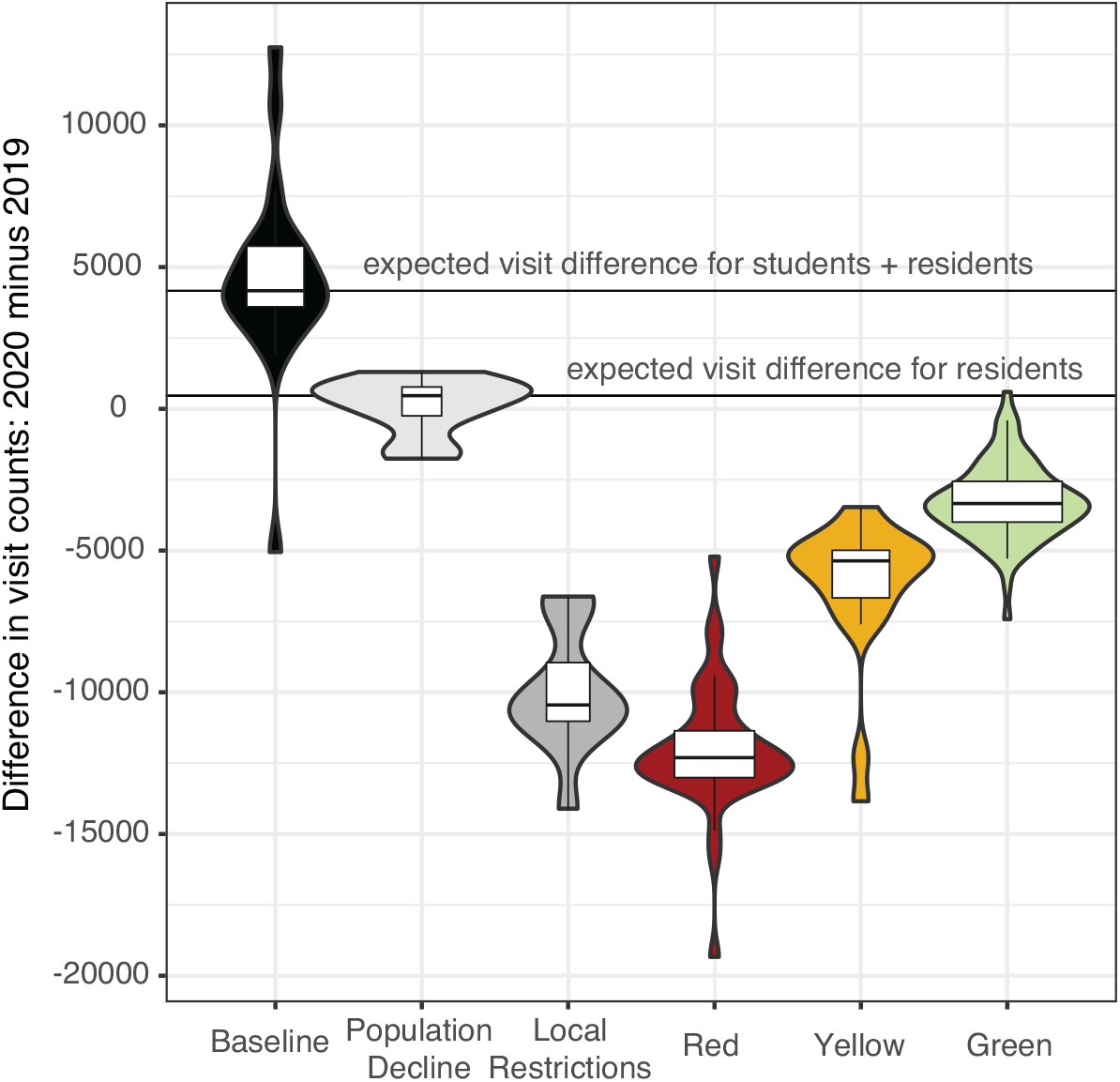
Aggregated mobile device visit counts. Summary of daily differences in visit counts between 2020 and 2019 within each phase. The width of boxplots within violin plots corresponds to the total number of days in each phase: minimum 8 days in Local Restrictions, maximum 78 days in the Red phase. The two horizontal lines indicate the median expected visit differential for 2020 compared to 2019 for students and residents combined (4,167) and for full-time residents only (464).

The largest negative differential between 2019 and 2020 visit counts occurred during the Red phase, reflecting the combined effects of the strictest restrictions in Centre County and the 2020 deficit of the student population (Figure 3, Figure 4B). Visit counts increased, reducing visit differentials during the Yellow and Green phases of restrictions. However, 2020 summer visit counts never reached 2019 values or expected visit counts based on pre-pandemic behavior from 2020, indicating ongoing and sustained changes in movement as measured by mobile devices in Centre County.

**Figure 4.**
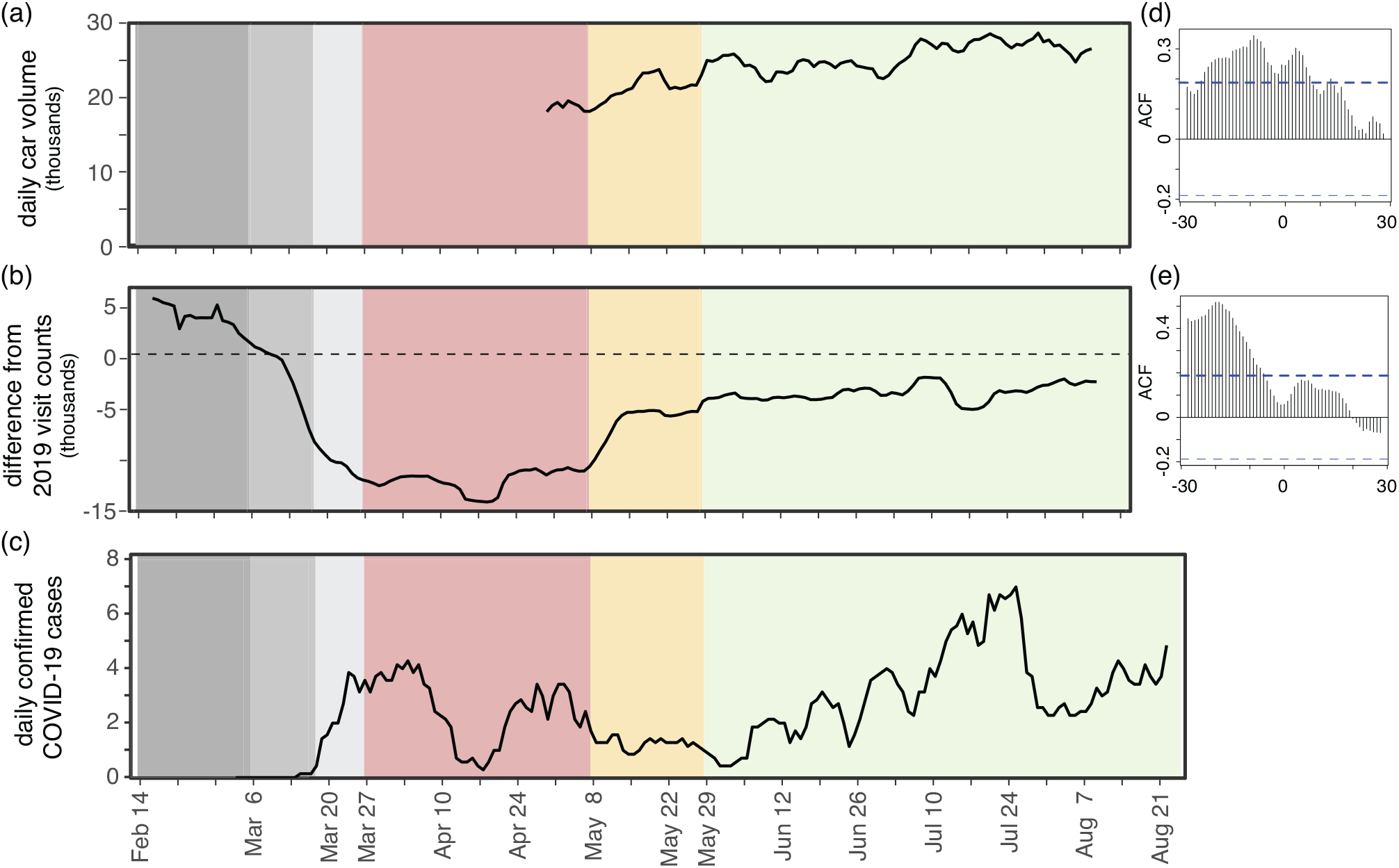
Estimated SARS-CoV-2 cases align with movement across phases. (A) 7-day rolling mean of daily total traffic volume increases as restriction phases ease. (B) 7-day rolling mean of daily differences in mobile device visit data between 2020 and 2019. (C) 7-day rolling mean of daily confirmed COVID-19 cases. (D) Autocorrelation function of daily traffic volume compared to 7-day mean of cases. Significant positive lags and leads are above the dotted blue line. (E) Autocorrelation function of visit differentials compared to 7-day mean of cases. Significant lags occur above the dotted blue line.

### Correlations between reported cases and movement

Daily COVID-19 case totals were low in Centre County throughout the summer of 2020 compared to urban and more populous counties in Pennsylvania [30]. Through 27 August 2020, Centre County confirmed a total of 448 cases, or 358.6 cases per 100,000 [30]. Daily incidence showed asynchronous outbreaks among neighboring counties in central Pennsylvania (Figure 1). In Centre County, increases in movement preceded reported cases (Figure 4). We find a significant correlation between traffic and a 7-day moving average of cases, with a 9-day lag in cases showing the greatest significance (ACF = 0.344, p <0.05; Fig 4a, d,). Additionally, we find a significant lagged correlation with mobile device visits to POI to reported cases. There is a significant lag from 28-to 7-days prior to cases, but the greatest correlation occurs at a 19-day lag (ACF = 0.518, p <0.05) (Figure 4b, e).

## Discussion

### Overview

Behavioral interventions were the primary large-scale public health tool available for preventing SARS-CoV-2 transmission throughout most of 2020. Behavioral interventions are most effective when implemented early and broadly. They are particularly important in areas where supportive care and medical resources are insufficient. Measuring levels of uptake of behavioral interventions is considerably more challenging than measuring uptake of pharmaceutical interventions but is critical for assessing current effectiveness and planning future outbreak response efforts. We used publicly available traffic volume and mobile device derived visits to POI to measure behavioral responses to COVID-19 restrictions in a rural county in central Pennsylvania. These data sources can be used to monitor movement, compliance with behavioral interventions, and adapt mitigation strategies in the future.

### Traffic and mobile devices

Data collected passively from traffic cameras measured movement in response to phased restriction policies. In Centre County, local roads provide access to essential businesses, which remained open throughout all restriction phases, while connector roads are used for travel between locations and may reflect movements to return to business, childcare and in-person work (i.e. during County Green phase). As restrictions eased, both local and connector vehicle volume increased, with greater increases on connector roads, particularly in the later Green phase. Although changes in restriction phases were announced approximately one week in advance, traffic volumes increased on the date that restrictions officially eased, and were not observed earlier. These results suggest that Centre County residents largely complied with county-level restrictions. However, this region did not transition from a less restrictive phase to a more restrictive phase during the period studied so compliance with enforcing stricter restriction phases cannot be assessed.

Daily SafeGraph mobile device derived visit counts and traffic volume showed similar patterns: movement gradually increased from the Red phase to the Yellow phase and Green phase. SafeGraph data were also critical in providing movement data from 2020 for dates prior to the implementation of restrictions phases as well as for corresponding dates from 2019. Data prior to the Red phase are not available for traffic volumes.

In the Red Phase of 2020, mobile device visit counts decreased by 81% compared to the baseline period of 2020. While some of this is due to the absence of students following spring break, these visit counts are still lower than visit counts from 2020 during pre-pandemic times when students were not in residence as well as the corresponding dates for 2019. The Green phase brought an increase in daily visit counts but the median daily visit count was still 11% lower than expected based on 2020 visit counts that preceded pandemic restrictions and lower than the corresponding time period in 2019. With SafeGraph’s longitudinal data, we were able to highlight compliance with restriction policies and calculate that visit counts in Centre County did not return to pre-pandemic levels during the summer of 2020, even as restrictions eased.

In Centre County, we collected data on more cars per day (mean for Red and Green phases) than mobile devices per day (mean for Red and Green phases), indicating the traffic data provide a better representation of the total county population than SafeGraph data. However, the differences in mobile device counts between the baseline and population decline periods of 2019 and 2020 show that SafeGraph data represent better students than residents. From March to August of 2019, the maximum mobile device visit counts from Centre County reflected known academic calendar events and university or community events near or on PSU’s UP campus (Figure S3).

### Movement and cases

Increases in movement following easing restrictions likely led to increased contacts and contributed to the subsequent uptick in cases during the green phase in Centre County. The bimodal pattern of infections in the summer most likely reflects increased SARS-CoV-2 transmission. We demonstrate a significant lag between movement measured by traffic and SafeGraph data and reported cases. Traffic data show significant lags and leads with reported COVID-19 cases. This is likely because the study period of the initial phase of the outbreak included increases in movement and cases and no decreases. SafeGraph data, which covered periods of declining and increasing movement only showed significant lags with cases. The significant lags between movement and cases highlight that movement underlies increases in transmission and precedes increases in reported cases by about two weeks. This provides an actionable metric for proactive policy, preventive actions, and modifying behavioral restrictions.

### Rural health

Many methods that are commonly used to track population movements are more effective in urban areas than in rural areas. Some methods rely on high smartphone usership and wireless network penetration or widespread and equitable access to high speed internet. Satellite based surveillance performs best on dense settlements with minimal cloud days. Due to high numbers of cloud days and small, sparse settlements, we could not measure these rural population dynamics with serial satellite imagery. However, we expect that any single data source cannot adequately represent a population, particularly a rural settlement. We intentionally used multiple, independently collected datasets to measure local movement patterns.

Urban population dynamics are often easier to measure that rural populations. Overall, urban populations also have greater access to health care, despite inequities within populations. They also have better-resourced, higher capacity health care centers when compared to rural areas throughout the United States, which improves pathogen surveillance and diagnostics. Rural populations experience vulnerabilities and uncertainties in health care that must be uniquely assessed and addressed. The COVID-19 pandemic has highlighted the necessity of locally responsive interventions. Due to reduced connectivity to populous locations, small towns experienced delayed SARS-CoV-2 introductions and lagged local outbreaks compared to urban centers [38]. Statewide regulations largely responded to urban outbreak fluctuations, which were not synchronized with rural outbreaks. When paired with effective federal response and relief, local oversight can most effectively serve outbreak response, management, and planning efforts.

## Conclusion

Early responses to the SARS-CoV-2 pandemic in the United States were managed by local governments implementing policies at state and county levels with insufficient resources or enforcement authority. COVID-19 spread widely through the nation’s big cities and small towns. Moving forward, it will be important to monitor local outbreaks and movement to design and implement locally responsive interventions. In addition to pharmaceutical interventions, measuring local population movements through passive approaches can help estimate uptake of behavioral interventions and adapt policies that target transmission prevention. Monitoring local movements and contacts is necessary to assess the impact of behavioral interventions intended to reduce pathogen transmission. Informed current and future uses of movement restrictions can help avoid overwhelming health care capacity, particularly in rural areas.

## Data Availability

Most data used for this study are available from publicly available sources and are attributed in the methods section. Academic researchers must register to receive access to SafeGraph data at no charge for non-commercial purposes only here: https://www.safegraph.com/academics. SafeGraph data are anonymized and aggregated. Our work with the anonymized and aggregated data provided by SafeGraph and used in the study does not qualify as human subjects research as defined by The Pennsylvania State University's IRB. SafeGraph approved the use of their data for this study. No additional data has been generated by this study. Code to process raw data, generate models and figures are available at: https://github.com/bhartilab/covid_movement_centre

## Supplementary material

The supplementary material for this article includes supplementary methods, figures S1-S3, and tables S1-S3.

## Acknowledgements

We thank the Pennsylvania Department of Transportation and the Pennsylvania Department of Health for providing publicly available data. We acknowledge financial support from Penn State University’s Huck Institutes of the Life Sciences and the Institute for Computational and Data Sciences. The funding sources had no role in the design, execution or interpretation of the results presented here.

## Ethics Statement

No ethical approval was required. The research used datasets that were freely available in the public domain. SafeGraph data are anonymized and aggregated.

## Data availability statement

All data used for this study are available from publicly available sources and are attributed in the methods section. No additional data has been generated by this study. Code to process raw data, generate models and figures are available at: https://github.com/bhartilab/covid_movement_centre

## SUPPLEMENTARY MATERIAL

### Supplementary Methods

#### I. Detailed timeline of events

Figure 1 in the main text details many of the key disease and policy events that are relevant to changes in policies and movement at the Penn State University Park campus and throughout Centre County. We provide more detail and citations for information in Figure 1 to give additional context to the policies and messaging from March through August of 2020.

**Timeline of important disease and policy events local study periods:**

Dec 31, 2019: China reports to WHO new pneumonia

Jan 20, 2020: 1st case in US

[BASELINE PERIOD: Feb 18 – March 6]

February 12: first death in US (Santa Clara, CA, retrospectively identified)[1]

March 6: first 2 reported cases of COVID-19 in PA

[LOCAL POPULATION DECLINE: Feb 1 – March 6]

March 9: PSU spring break begins[2]

March 11: WHO declares COVID-19 a pandemic[3]; PSU announces no resident instruction following spring break[4]

March 16: PSU begins remote instruction

March 19: all non-life-sustaining businesses ordered to close statewide by PA Governor[5]

March 21: enforcement of closure of non-life-sustaining businesses statewide[5]

[RED PHASE: Feb 1 – March 6]

March 28: Centre County receives stay at home order from PA Governor [6]

April 3: PA Governor calls for “universal masking”

April 9: PA schools officially closed through end of academic year

May 1: announcement from PA Governor that on May 8 PA will lift some restrictions in 24 counties in the Northcentral and Northwest health districts; all counties will remain in “red” until May 8, these counties will move to “yellow” on May 8 [6]

[YELLOW PHASE: Feb 1 – March 6]

May 8: PA lifts some restrictions in 24 counties in the Northcentral and Northwest health districts as determined by PA Governor in moving from “red” (Stay-at-home) to “yellow” (aggressive mitigation) [6]

May 28: US death toll from COVID-19 passes 100,000

[GREEN PHASE: May 29 – August 14]

May 29: Some of Northcentral and Northwest health districts (Fig S2) move from “yellow” to “green”

June 30: First PSU student death from COVID-19

July 1: Order of face coverings in public places in Pennsylvania [7]

#### II. Details of traffic camera acquisition, standardization, and cleaning

We captured images every 20 seconds from each of 19 cameras. To standardize variation in images captured per hour, with variation due to timing mismatches or missing images (see below), the sum of vehicles counted each hour is divided by the number of images captured each hour and scaled by the number of expected images if we captured one image per minute. This avoids overcounting cars at traffic lights but likely underestimates total vehicle volume on interstates. Therefore, count data should be interpreted as relative increases rather than absolute.

Occasionally, live stream traffic cameras would fail to capture images correctly, resulting in missing images. The standardized hourly counts were cleaned to remove strings of zeros or integers, which were the result of frozen images or occasionally, parked cars. None of the cameras capture legal parking zones when they are in the correct position. Incorrect vehicle counts from missing or erroneous images were replaced with NAs and predicted with the best fit generalized additive model (see Methods in Main Text; Fig S2).

## Supplementary Tables

**Table S1.** Traffic camera locations and descriptions.

| ID | Latitude | Longitude | Ownership | Number of lanes | Road Type | Description |
| --- | --- | --- | --- | --- | --- | --- |
| CAM02001CCTV2 | 40.810961 | -78.075259 | PennDOT<br>(Pennsylvania Department of Transportation) | 5 | connector | Port Matilda US-322 By-Pass Westbound |
| CAM02002CCTV3 | 40.817144 | -77.939841 | PennDOT | 7 | connector | Grays Woods |
| CAM02003CCTV4 | 40.828613 | -77.840339 | PennDOT | 6 | connector | I-99/US-322 Interchange |
| CAM02005CCTV9 | 40.955477 | -77.773749 | PennDOT | 3 | connector | Milesburg Interchange West I-80 Exit 158 Eastbound / Alt US-220 |
| CAM02006CCTV10 | 40.956061 | -77.766277 | PennDOT | 3 | connector | Milesburg Interchange East I-80 Exit 158 Westbound / PA-150 |
| CAM02007CCTV13 | 40.795773 | -77.820937 | PennDOT | 4 | connector | US-322 E/O PA-26 |
| CAM02009CCTV7 | 40.944571 | -77.720918 | PennDOT | 2 | connector | I-80 Eastbound Exit 161 Bellefonte Interchange (I-80 & I-99) |
| CAM02010CCTV11 | 40.80388 | -78.063664 | PennDOT | 6 | connector | Port Matilda I-99 Exit 61 Median |
| CAM02020CCTV24 | 40.829048 | -77.804817 | PennDOT | 6 | internal | Benner Pike |
| CAM02028CCTV32 | 40.812073 | -77.9225 | PennDOT | 6 | connector | Atherton and Valley Vista |
| CAM02033CCTV38 | 41.022703 | -77.933981 | PennDOT | 4 | connector | Snowshoe I-80 Exit 147 |
| CAM02037CCTV43 | 40.807936 | -77.895204 | PennDOT | 5 | internal | Atherton and Vairo |
| CAM02038CCTV44 | 40.805031 | -77.886886 | PennDOT | 5 | internal | Atherton and North Hills |
| CAM02039CCTV45 | 40.796522 | -77.872549 | PennDOT | 5 | internal | Atherton and Park |
| CAM02040CCTV46 | 40.791787 | -77.864971 | PennDOT | 4 | internal | Atherton and W.College |
| CAM02042CCTV49 | 40.790894 | -77.863812 | PennDOT | 4 | internal | Atherton and W.Beaver |
| CAM02046CCTV52 | 40.817953 | -77.900271 | PennDOT | 4 | connector | I-99 at Exit 71 (Toftrees) |
| collegeTwp | 40.811564 | -77.830719 | State College Township | 4 | connector | College Township Traffic camera |
| parkArboretum | 40.804483 | -77.864219 | Penn State University | 3 | internal | Penn State University Park Ave. |

**Table S2.** Calendar dates for phases between 2020 and 2019 for SafeGraph data. The timing of Spring Break and Commencement at Penn State determine large scale movement events in State College and surrounding areas in Centre County. The timing of these events was matched across 2019 and 2020 so events during the pandemic year would be accurately compared to the same events/timing in previous year. All phases have the same number of days and number of weekdays. For the Population Decline period, the dates included the Friday before spring break through the last Sunday of spring break. Dates are from the Office of the Registrar at Penn State [2].

| Year | Baseline | Population Decline (Spring Break) | Local Restrictions | Red | Yellow (corresponds to commencement) | Green |
| --- | --- | --- | --- | --- | --- | --- |
| 2019 | Feb 8 | March 1 | March 11 | March 22 | May 3 | May 24 |
| 2020 | Feb 14 | March 6 | March 18 | March 27 | May 8 | May 29 |

**Table S3.** Summary of daily traffic data. Estimates of daily means of vehicle counts using observed and predicted counts from best-fit GAM.

|  |  | **Mean daily traffic by Restriction Phase** |  |  |
| --- | --- | --- | --- | --- |
| **Weekday/**<br>**Weekend** | **Road type** | **County Red** | **County Yellow** | **County Green** |
| Weekday | Internal | 10772 | 13267 | 15309 |
| Weekday | Connector | 7397 | 9729 | 12425 |
| Weekend | Internal | 12678 | 10455 | 12083 |
| Weekend | Connector | 6367 | 6367 | 8113 |

## Supplementary Figures

**Figure S1.**
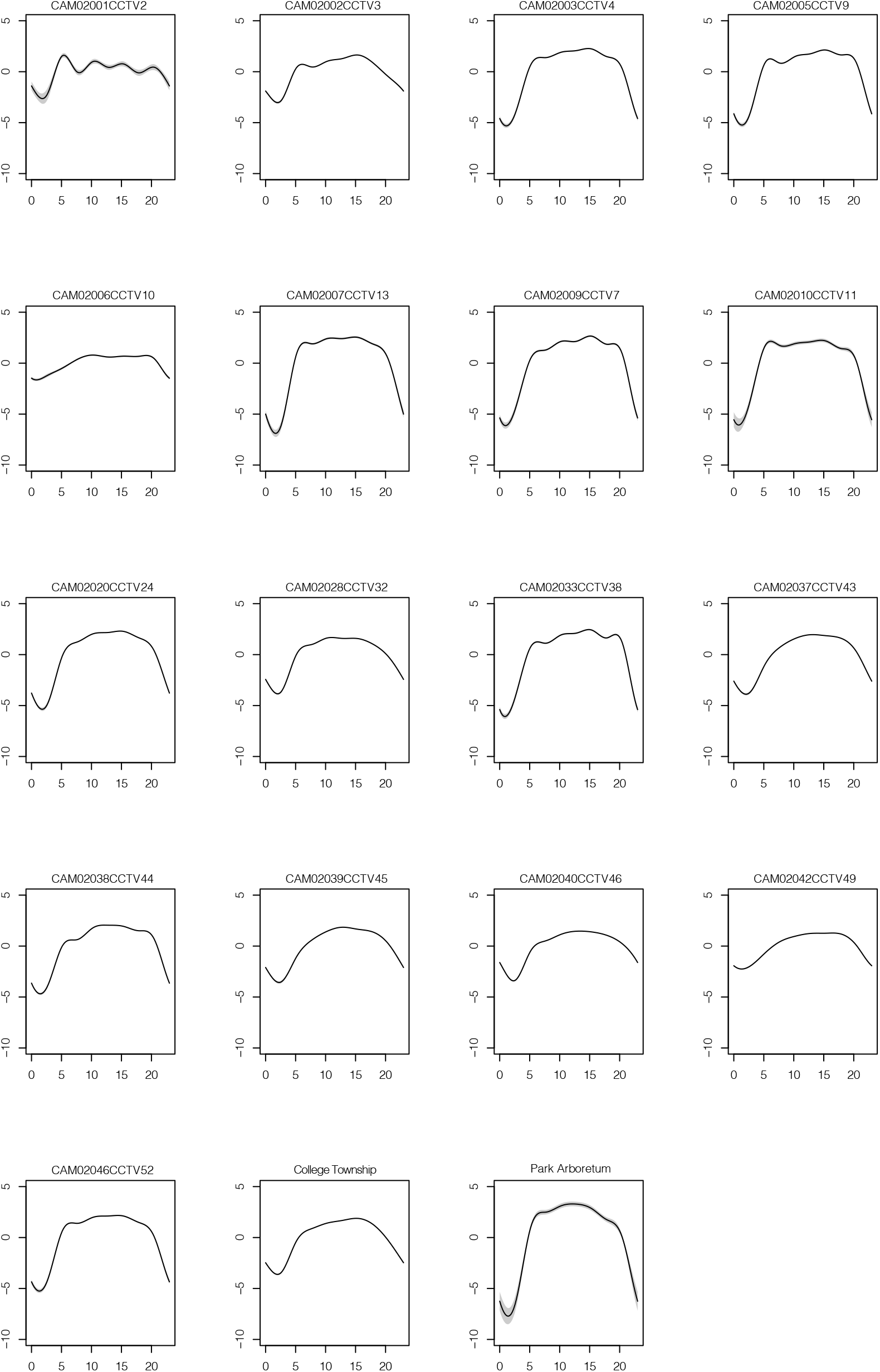
Splines fit to hourly traffic volume by individual cameras. The y-axes show the estimates of the best-fit GAM and x-axes show the hour of the day (range 0-23).

**Figure S2.**
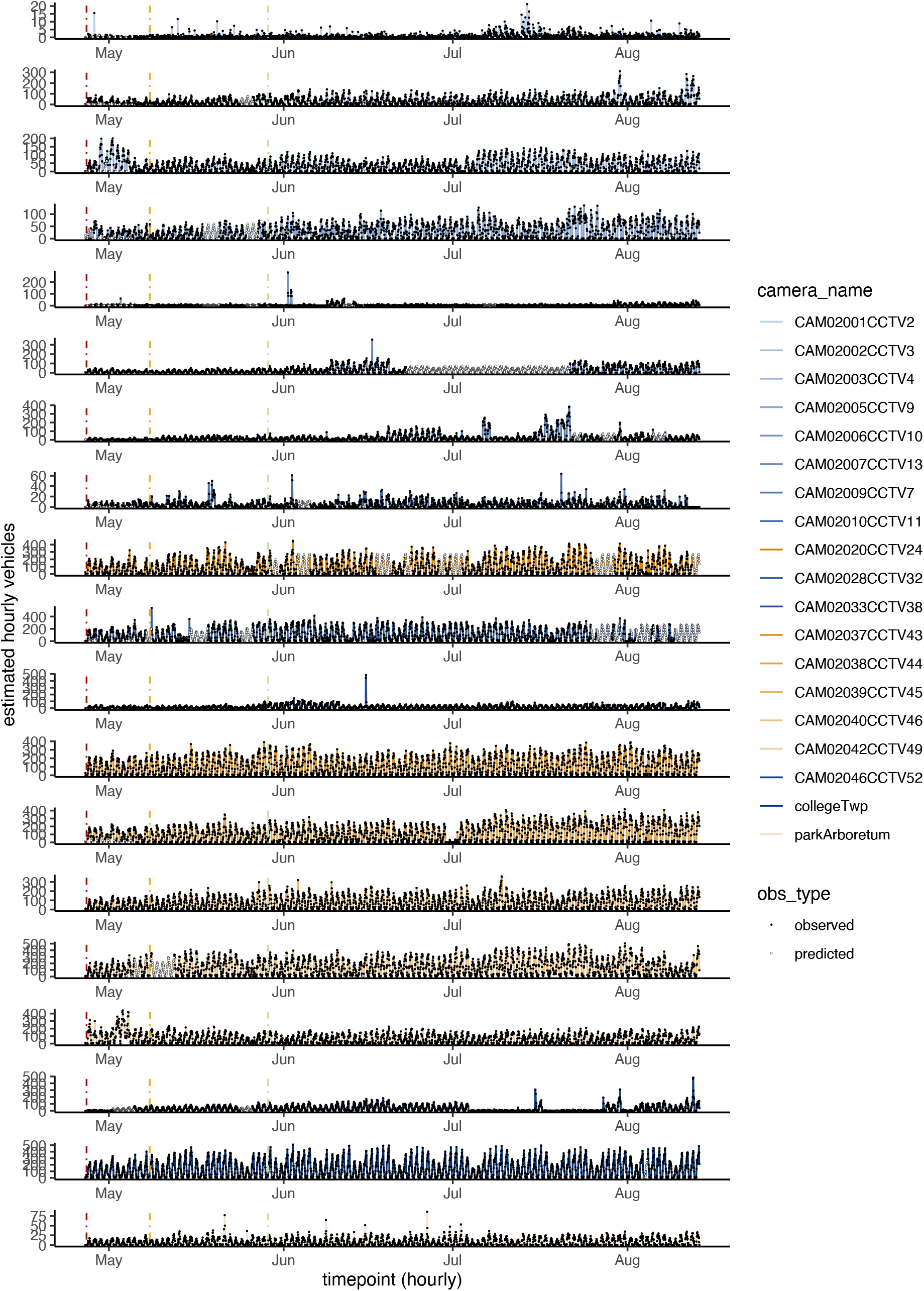
Observed and predicted hourly count data per camera through time. Predicted counts from the best-fit GAM are used to fill in gaps in observed data and are shown in open circles. Phases are indicated with vertical colored lines for Red, Yellow and Green Phases. Colors for each camera are consistent with Fig 2 in the main text; shades of blue represent connector roads and orange shades represent internal roads. The range of the y-axis varies by camera.

**Figure S3.**
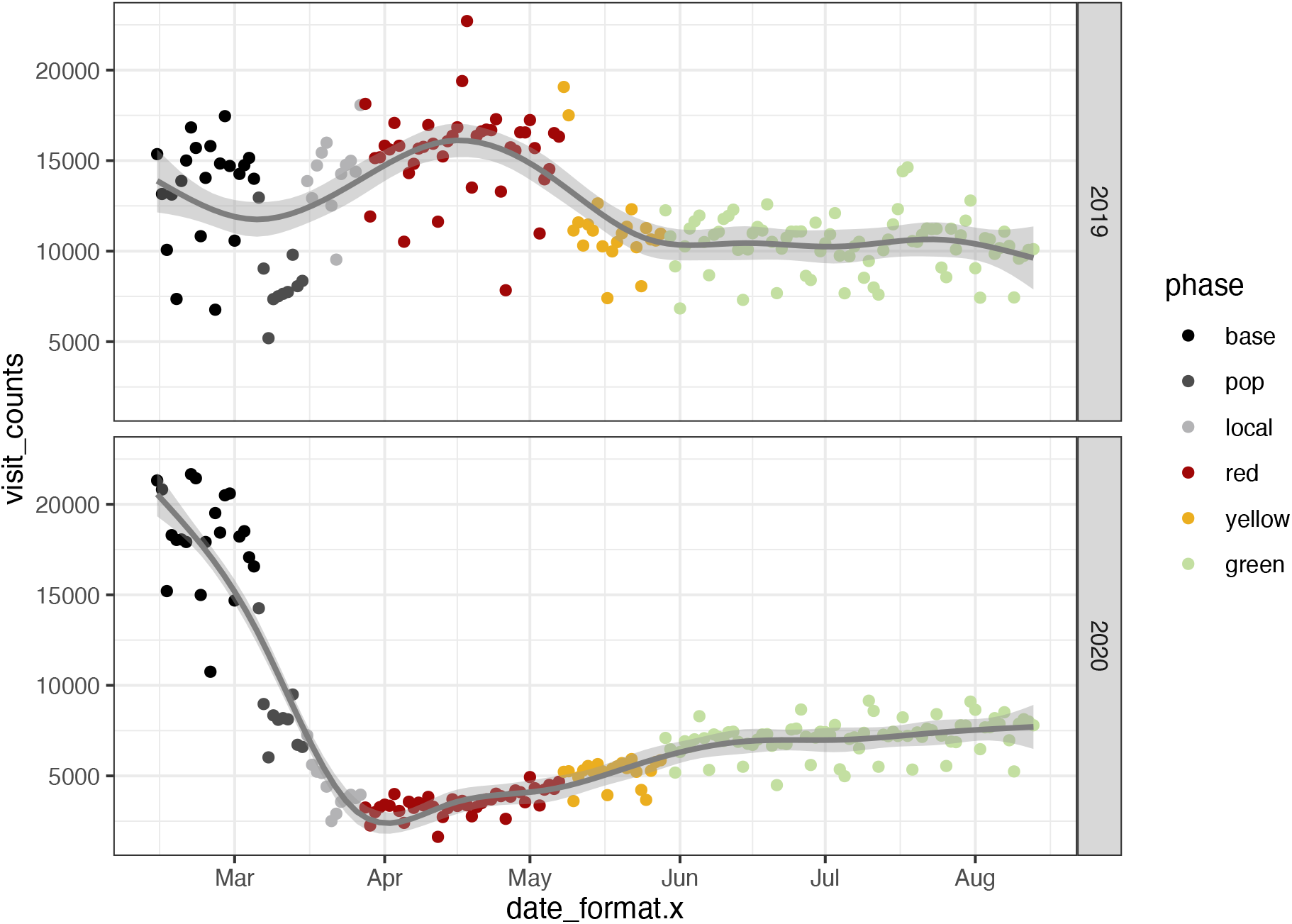
Visit counts for Centre County, Spring and Summer 2019-2020.

